# Why context matters: Clinical validation of digital measures for use in Parkinson’s disease

**DOI:** 10.1101/2025.10.14.25337984

**Authors:** Anthony D Scotina, Jennie S Lavine, Seth Haney, Jessie P Bakker, John A. Wagner, Larsson Omberg

## Abstract

The clinical validation of digital measures for Parkinson’s disease (PD) must be tailored to their specific purpose, or context of use (CoU). However, validation work often assumes that the same measures will work across contexts. We analyzed data from the ObjectivePD substudy of the mPower study, involving 24 people with PD (PwPD) and 22 healthy controls (HCs). We evaluated 31 digital measures from five smartphone-based assessments (gait, tapping, and three types of tremor) for test-retest reliability, criterion validity against the MDS-UPDRS, and performance across three CoUs: differentiating PwPD from HCs, detecting effect of symptomatic treatments, and tracking disease progression. Of the 31 measures, 22 were found to be reliable (Intraclass Correlation Coefficient >0.5), and many correlated significantly with MDS-UPDRS items, such as postural tremor root mean squared (RMS) acceleration (τ = 0.51) and finger tapping speed (τ = -0.51). Performance was context-specific, however: certain measures excelled at differentiating PwPD from HCs, others were highly sensitive to medication effects, and a different set tracked long-term changes. The validity of a digital measure is not universal but depends on its intended application. This context-driven approach is essential for establishing that a measure is fit-for-purpose and for qualifying its use in future PD clinical trials.

## Introduction

Validation of measures derived from a sensor-based digital health technology (sDHT) requires multiple levels of evidence to demonstrate that they are fit-for-purpose, the V3 and V3+ framework categorizes the verification into the following: sensor verification, usability validation, analytical validation, and clinical validation^1,2^, the latter of which is highly dependent on the proposed context of use (CoU).^3^ A growing body of work has evaluated various aspects of clinical validity of sDHT-derived measures in Parkinson’s disease (PD); for example, Lipsmeier et al found that several measures from the Roche-PD mobile app (F Hoffman-La Roche; Basel, Switzerland) demonstrated moderate to excellent test–retest reliability, known-groups validity distinguishing between people with Parkinson’s disease (PwPD) and healthy controls (HCs), and criterion validity against corresponding clinician-reported Movement Disorder Society-Unified Parkinson’s Disease Rating Scale (MDS-UPDRS) items.^4^ The ObjectivePD substudy of the mPower study reported that measures derived from a smartphone-based finger tapping assessment demonstrated known-groups validity and criterion validity.^5^ Similarly, Adams et al. found that sDHT-derived measures of gait, tremor, and speech captured in the *Wearable Assessment in The Clinic and at-Home in PD* (WATCH-PD) study demonstrated criterion validity at baseline, though responsiveness to change over 12-months was only demonstrated for assessments administered in-clinic and not for those administered at-home.^6,7^ Such findings support criterion validity of digital measures by linking them to “gold-standard” assessments (MDS-UPDRS items) and suggest these tools show some degree of responsiveness to long-term changes in symptoms. Additionally, studies have shown that continuous monitoring can detect symptom fluctuations and treatment effects that sporadic in-clinic exams may miss; for instance, sDHT-based assessments have captured short-term improvements with levodopa and other symptomatic treatments in real-time. ^4,5,8^

Research to date has largely focused on evaluating the clinical validity of digital measures without clearly defining the CoU, despite its centrality in the regulatory environment for qualification of digital biomarkers.^9,10^ Clinical validity, defined as evidence that the digital measure acceptably identifies, measures, or predicts a meaningful state or experience,^1^ is typically evaluated using the concepts of reliability and validity borrowed from the field of psychometrics. Notably, the BEST (Biomarkers, EndpointS, and other Tools) resource co-developed by the United States Food and Drug Administration (FDA) and National Institutes of Health differentiates seven distinct biomarker categories (susceptibility/risk; diagnostic; monitoring; prognostic; predictive; pharmacodynamic/response; and safety; ^11^) and expects validation data appropriate to each. A one-size-fits-all approach to clinical validation is therefore insufficient; instead, developers must tailor their research and development strategy to a specific CoU by describing the way the digital measure is to be used and the purpose of the use, including the intended population(s) of interest and the intended use environment(s). For example, many studies have not specified whether a given digital measure was being validated as a diagnostic, monitoring, or response biomarker, ^12^ which is increasingly recognized as an important gap in the field. As a result, it remains unclear if certain digital measures are fit-for-purpose for all use cases or if their performance varies by application. A digital biomarker intended as a PD diagnostic tool, for example, would need to demonstrate high sensitivity and specificity in distinguishing patient groups; essentially, strong known-groups validity. In contrast, a measure applied as a monitoring biomarker in the context of a clinical trial endpoint must show longitudinal responsiveness to change as well as construct validity demonstrating that the measure adequately reflects disease severity. ^4^

In the present secondary analysis of the ObjectivePD substudy of mPower, ^5,13^ we have evaluated the reliability and clinical validity relevant to the use of a toolkit of digital measures of PD motor symptoms derived from five active assessments performed on a smartphone without supervision both at-home and in-clinic. This rigorous, context-driven approach to clinical validation is not only scientifically sound but also aligned with regulatory expectations, facilitating future acceptance and/or qualification of digital measures in PD trials. We then considered three specific applications and determined which measures show evidence of being fit-for-purpose in each: (1) does the measure differentiate between healthy adults and those with early PD (required for example for a diagnostic biomarker); (2) does the measure respond to short-term fluctuations associated with symptomatic treatment medication (required for example for a pharmacodynamic/response biomarker); (3) does the measure respond to long-term changes in progression of PD symptoms (required for example for a monitoring biomarker)? Finally, we discuss the relevance of our findings for the development of a holistic digital toolkit that can support upcoming trials of PD disease-modifying therapies.

## Methods

### Study description

Data used in the current analyses were generated by processing raw sensor data from the ObjectivePD substudy of the mPower study, in which 24 PwPD and 22 HCs were enrolled for at least 6-months to complete daily mobile app-based active assessments in the home setting and in-clinic visits at 0, 3, and 6 months.^5^ Digital assessments included (1) 30 seconds of speeded finger tapping alternating between the index and middle finger, (2) 30 seconds of gait with the smartphone in the pocket, and (3) resting, postural, and hand-to-nose (kinetic) tremor assessments all of which were done for 10 seconds. PwPD completed assessments in both the ON and OFF state; all analyses described here used data captured in the OFF state unless otherwise specified.

This study used publicly available, pseudonymized data and is not considered human subjects research under the common rule in the United States and is being used in accordance with the governance rules of the data as defined at: https://www.synapse.org/Synapse:syn4993293/wiki/247860.

### Data processing

Raw accelerometer and finger tapping (touchscreen) event data were processed by algorithms developed at Koneksa Health (New York, NY, USA) to generate digital measures listed in **Supplementary Table 1** and described previously.^14,15^ Measures captured during assessments that were completed out of compliance with the study protocol were removed; specifically, finger tapping assessments were removed if ≤5 taps were recorded; gait assessments were removed if the duration was <18 or >35 seconds; resting and postural tremor assessments were removed if the duration was <8 seconds; and kinetic tremor assessments were removed if there were <4 hand-to-nose touches. All assessments that yielded measurements >3 standard deviations away from the mean were manually reviewed for potential removal due to compliance issues.

### Statistical methods

We calculated test-retest reliability using intraclass correlation coefficients (ICCs) derived from linear random intercept models, as described previously.^15^ Specifically, digital measurements were grouped per 21 days, and a linear model was fit per measure with random intercepts for participant and participant-by-group interaction. This procedure was used to estimate the test-retest reliability separately for measures derived from individual at-home assessments and for measures derived from multiple at-home assessments completed during a 7-day window (that is, a “burst”). For the latter, we treated the median of measures derived from the burst as the dependent variable in each linear model.

Criterion validity was assessed by comparing daily at-home digital measures with aligned in-clinic MDS-UPDRS Part 2 and Part 3 items; for example, tremor frequency for left hand postural tremor was compared with MDS-UPDRS item 3.15 *Postural Tremor of the Hands* (see **Supplementary Table 1** for a complete list of comparisons). Because at-home digital measures and in-clinic MDS-UPDRS were collected at different timepoints, we retained at-home measures that were collected within 10 days of an in-clinic visit, excluding digital measures derived from assessments that were within two hours of a visit (to avoid erroneously including digital assessments performed under possible supervision in the clinic). Within each 20-day window around an in-clinic visit, we aggregated daily at-home digital measures by calculating the median. We then assessed the relationships between digital measures and MDS-UPDRS items using Kendall’s tau (for MDS-UPDRS items scored on a Likert scale) or Pearson’s correlation (for the MDS-UPDRS tremor subscore; that is, the sum of items 3.15–3.18 and 2.10). To isolate any effect of time we evaluated criterion validity at baseline as well by repeating these analyses comparing Day 0 MDS-UPDRS scores to the median of at-home digital measures captured during the first 10 days in the study. Finally, we compared digital measures derived from assessments completed during in-clinic visits to MDS-UPDRS items, using data from all three visits (see **Supplementary Table 2**).

We used linear mixed-effects models (LMMs) to evaluate known-groups validity and responsiveness to change. To compare measures on a uniform scale, we scaled each by their minimum detectable change (MDC)^16^ by first extracting the residual variance from each LMM (σ_*m*_^2^) and then calculating MDC as follows: 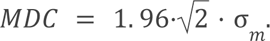 Each set of models specified each respective digital measure as the dependent variable and included a participant-specific random intercept. Three different modeling approaches were then performed: A) known-groups validity distinguishing PwPD from HCs with fixed effects for time and group, in which effect size is defined as the estimated MDC-scaled difference between groups; B) responsiveness to short-term change distinguishing ON versus OFF state amongst PwPD taking medication, with fixed effects for time (continuous days from baseline) and medication timing (before versus after; that is, ON versus OFF state), in which effect size is defined as the estimated MDC-scaled difference between OFF and ON states; and C) responsiveness to long-term change as measured by differential progression rates of PwPD versus HCs with fixed effect for the time-by-cohort interaction, in which effect size is defined as the difference in estimated MDC-scaled change per year between PwPD and HCs. Differential progression rate instead of progression rate in the PwPD population was chosen to isolate disease specific progression from other changes such as those due to practice effects, participants becoming complacent as a study progresses or natural progression unrelated to the disease. This was in part motivated by prior work that showed that finger tapping speed improved in PwPD^7^ as well as in HCs.^7,17^

Within each analysis domain, the Benjamini-Hochberg method was used to adjust for multiplicity using a false-discovery rate of 0.05. ^18^ We performed all data processing and analysis using Python 3.12.

## Results

### Study participants and data quality

Twenty-four PwPD and 22 HCs participated in ObjectivePD (see **Table 1**). On average, PwPD were older and more likely to be men, compared to HCs. The PwPD cohort tended to optionally participate beyond the 6-month timeframe and contributed on average 1,400 more smartphone-based assessments per participant than the HC cohort (2,015 versus 615, respectively).

**Table 1:**
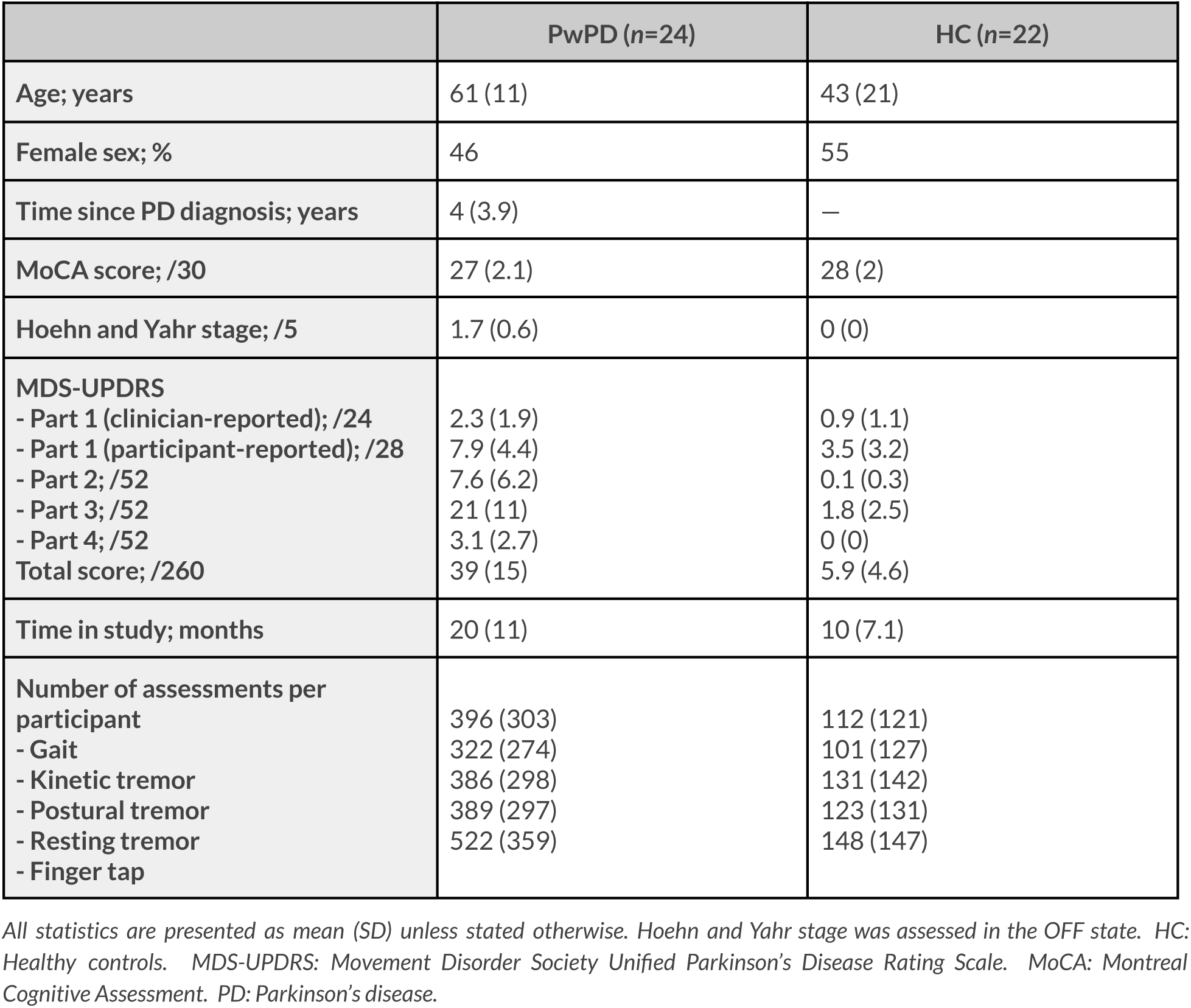
Baseline characteristics of the ObjectivePD study population.

Of the 32,056 total contributed assessments, 1,861 (5.8%) were removed due to evidence that they were completed out of compliance with the assessment instructions. Specifically, 0 of the tapping assessments, 550 (8.2%) of gait assessments, 135 (1.2%) of resting tremor assessments, 39 (0.35%) of postural tremor assessments, and 1,137 (10%) of kinetic tremor assessments were filtered out. Only two assessments, both kinetic tremor (<0.02%) were removed because they were >16 standard deviations from the mean.

From the five assessment categories, we generated 31 individual digital measures (13 for gait; 4 each for resting, postural, and kinetic tremor; and 6 for finger tapping (see **Supplementary Figure 1** and **Supplementary Table 1**).

### Test-retest reliability

Of the 31 digital measures, 27 and 22 exhibited moderate to excellent test-retest reliability (ICC >0.5)^19^ when evaluated across 7-day bursts and across individual assessments, respectively (see **Figure 1**). All gait measures exhibited moderate to excellent reliability when compared across bursts, and all but step time discrepancy did when compared across individual assessments. Several measures showed poor reliability across both measurement schedules; for example, kinetic tremor peak frequency of displacement (ICC 0.43 across bursts and 0.20 across individual assessments) and finger tap interval change (ICC 0.25 across bursts and 0.09 across individual assessments).

**Figure 1:**
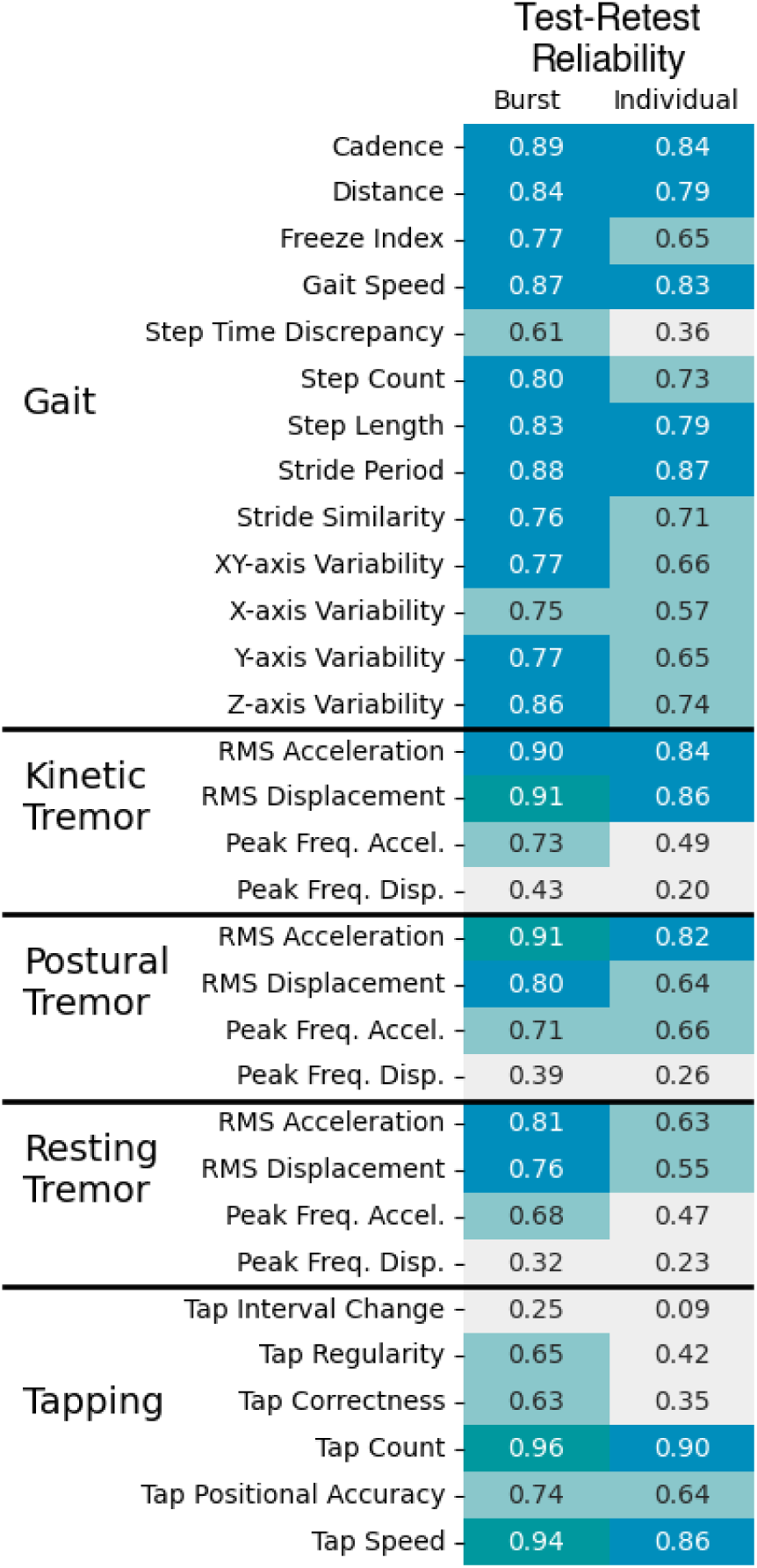
Test-retest reliability of digital measures evaluated across bursts and individual assessments. *Cells are color-coded according to ICC thresholds: dark green for excellent (ICC >0.9), turquoise for good (ICC >0.75), and light turquoise for moderate (ICC >0.5).*

### Criterion validity

To evaluate criterion validity, each digital measure was aligned with at least one item each from MDS-UPDRS Part 2 and Part 3; tremor features were also compared against the tremor subscore (see **Supplementary Table 1**). Of the 31 digital measures, 12 and 17 demonstrated a statistically-significant correlation with items from MDS-UPDRS Part 2 and Part 3, respectively, while 7 of the 12 measures derived from tremor assessments demonstrated a statistically-significant correlation with the tremor subscore (see **Figure 2**). The strongest observed relationships were between postural tremor root mean squared (RMS) acceleration and MDS-UPDRS item 3.15 (*Postural Tremor of the Hand*s; tau = 0.51) and between finger tap speed and item 3.4 (*Finger Tapping*; tau = -0.51). Each postural and resting tremor RMS acceleration and displacement measure demonstrated a statistically-significant correlation with its aligned MDS-UPDRS item (2.10 *Tremor*; 3.15 *Postural Tremor of the Hands*, or 3.17 *Rest Tremor Amplitude*). Each finger tap measure demonstrated a statistically significant correlation with item 3.4 (*Finger Tapping*); only tap count demonstrated a statistically-significant correlation with item 2.5 (*Dressing*).

**Figure 2:**
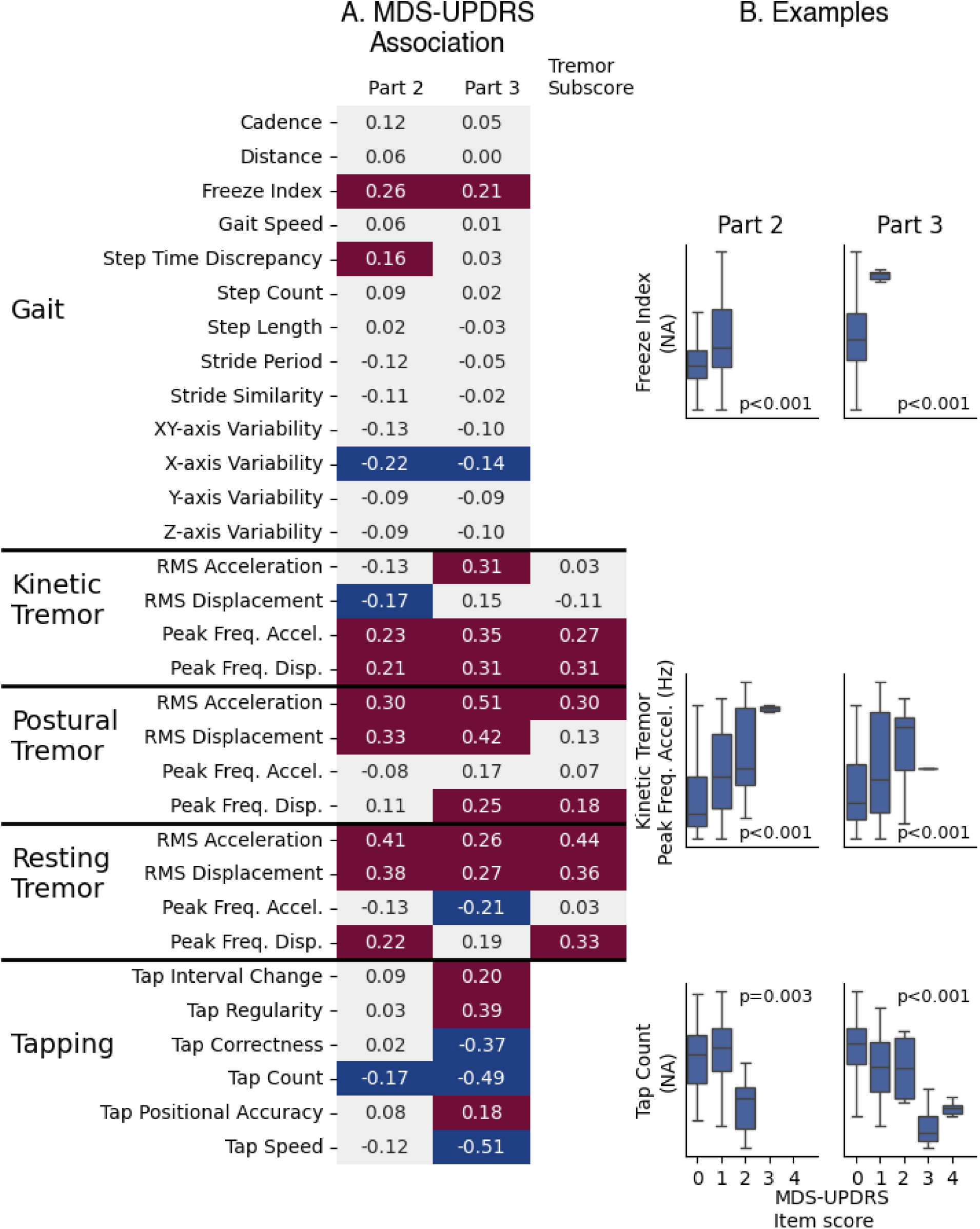
Criterion validity comparing digital measures to aligned MDS-UPDRS items. *A) depicts the effect size for associations between digital measures and aligned MDS-UPDRS items. Each tremor and finger-tapping measure was compared against more than one item from MDS-UPDRS Part 3 (see **Supplementary Table 1**); the effect sizes shown here are those with the lowest p-value. Red cells and blue cells indicate statistically-significant positive and negative associations, respectively, following adjustment for multiplicity. B) An example for one measure per assessment, with p-values based on tests depicted on the left. The Freeze Index is shown compared with MDS-UPDRS items 2.13 (Freezing) and 3.11 (Freezing of Gait). Peak Frequency of Acceleration from Kinetic Tremor is shown compared with items 2.10 (Tremor) and 3.16a (Kinetic Tremor of the Hands; right). Tap Count is shown compared with items 2.5 (Dressing) and 3.4b (Finger Tapping; left). Effects sizes shown here were computed using either Kendall’s tau (for evaluations of individual MDS-UPDRS items) or Pearson’s correlation (for evaluation of the MDS-UPDRS tremor subscore)*.

In addition to evaluating criterion validity using MDS-UPDRS scores captured during all three in-clinic visits, we analyzed data captured only during the first month of the study (that is, baseline data). We found that in this smaller database a subset of 11 measures which exhibited statistically-significant criterion validity in the full dataset also exhibited baseline criterion validity (see **Supplementary Table 2**), likely due to the smaller sample size of a single study month. Finally, the criterion validity for measures collected in-clinic and not at-home were similar with a tendency for the correlation to be higher in-clinic compared to at-home. There were a few additional associations, such as stride similarity versus MDS-UPDRS item 2.12 (*Walking and Balance*; tau = -0.56), and postural tremor RMS displacement versus the MDS-UPDRS tremor subscore (tau = 0.49) but these could be due to the group FDR rate (see **Supplementary Table 2**).

### Known-groups validity

We assessed known-groups validity by comparing digital measures from PwPD versus HCs while controlling for time elapsed (in days) since the start of the study. Of 31 measures, 6 were able to statistically differentiate between the two groups (**Figure 3**), with tap positional accuracy and postural tremor peak frequency acceleration showing the largest observed effects (-0.44 and 0.41, respectively). Of note, none of the gait assessment measures were able to statistically differentiate between these known-groups.

**Figure 3:**
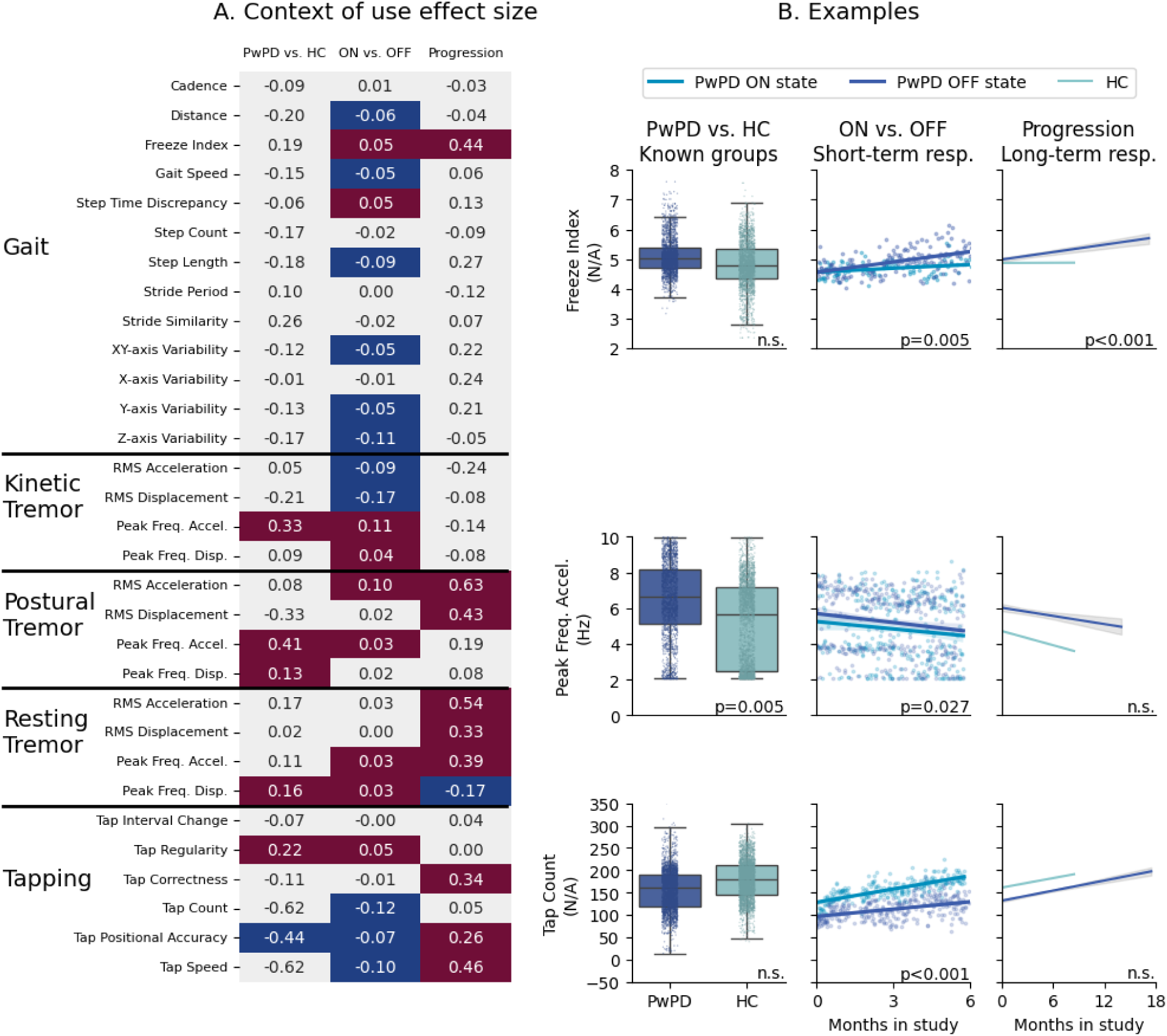
Performance of digital measures according to known-groups validity and responsiveness to short-term and long-term change. A) *Effect sizes, for 1) known-groups validity; that is, differentiating PwPD and HC cohorts; 2) responsiveness to short-term change; that is, differentiating ON versus OFF state; and 3) responsiveness to long-term change; that is, differential progression of PwPD and HC cohorts. Red cells and blue cells indicate statistically-significant positive and negative associations, respectively, following adjustment for multiplicity. B) Example for one measure per assessment, with p-values based on tests depicted in Panel A. The ON versus OFF state figures contain data from a single participant while figures depicting responsiveness to short-term and long-term change are based on all available data and model parameters respectively*.

### Responsiveness to short-term change

In data from PwPD, we assessed the impact of medication state (ON vs. OFF state) to evaluate responsiveness to short-term change. Twenty measures showed a significant association with medication state, with each assessment contributing at least two significant measures (**Figure 3).** RMS displacement of kinetic tremor showed the largest effect (-0.17).

### Responsiveness to long-term change

Ten measures from all assessments except kinetic tremor showed significant differential progression between PwPD and HC, thereby exhibiting long-term responsiveness to change. **(Figure 3)**. These included items such as *Freeze Index* (0.44) from gait, the RMS Acceleration in postural tremor (0.63) and in resting tremor (0.54). Of note, three measures from the finger tap assessment exhibited differential progression where the rate of progression was unexpectedly higher for PwPD compared with HCs; for example, finger tap speed increased over time on average amongst PwPD, and increased at a faster rate compared to HC. While finger tap count did not demonstrate differential progression between PwPD and HC, it increased over time for both cohorts.

### Performance of digital measures across clinical validity domains

The results described above emphasize that different digital measures exhibit measurement properties relevant to different aspects of clinical validity. For example, *Freeze Index* captured during gait assessments was shown to be A) reliable when evaluated across bursts (ICC 0.77) and individual assessments (ICC 0.65); B) significantly associated with MDS-UPDRS items 2.13 (patient-reported *Freezing*; p<0.001) and 3.11 (clinician-reported *Freezing*; p<0.001); C) responsive to short-term change by differentiating between ON versus OFF state (0.05 MDC-scaled units, or 0.06 units, less in the ON state; *p*=0.005); and D) responsive to long-term change by detecting differential disease progression between PwPD and HCs (0.44 MDC-scaled units, or 0.49 units, more per year in PwPD; *p*<0.001). However, *Freeze Index* was not able to statistically differentiate between PwPD and HC at a fixed point in time. **Figure 4** summarizes all evaluated digital measures demonstrating at least moderate test-retest reliability (ICC ≥0.5), categorized according to performance across three domains of clinical validity (known-groups validity; responsiveness to short-term change; responsiveness to long-term change).

**Figure 4:**
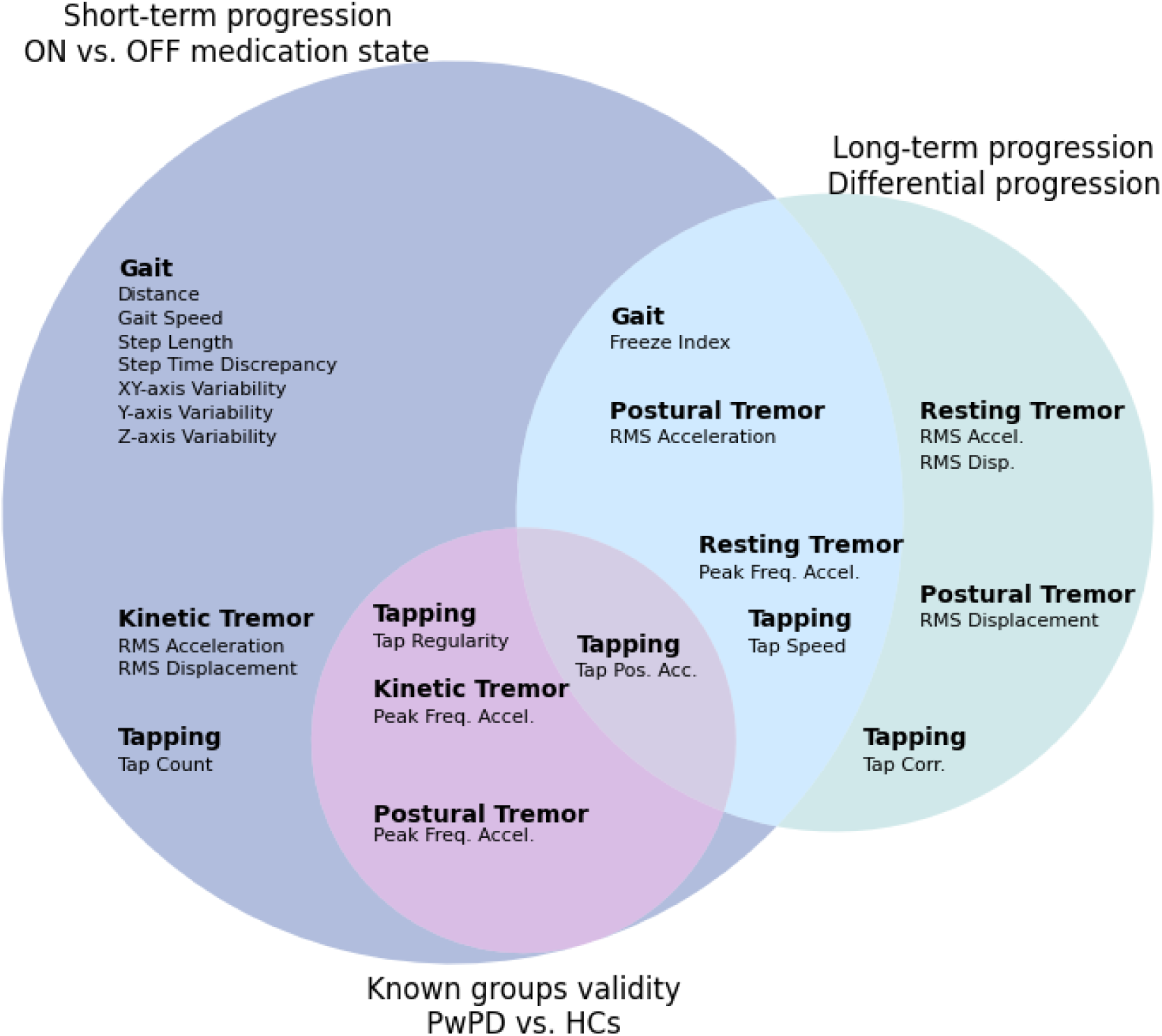
Reliable digital measures categorized according to three domains of clinical validity. *Measures depicted in this Venn diagram are those demonstrating moderate to excellent test-retest reliability (that is, excluding measures with ICC <0.5) and statistically significant results for at least one clinical validity domain (known-groups validity differentiating PwPD and HCs; responsiveness to short-term change differentiating ON versus OFF medication state; and responsiveness to long-term change as measured by differential progression rates of PwPD versus HCs).*

## Discussion

In this paper, we evaluated the reliability and clinical validity of 31 digital measures of PD motor symptoms derived from five smartphone-based assessments performed at-home without supervision, and found that 22 measures (9 from tremor, 8 from gait, and 5 from finger tapping) exhibited moderate to excellent test-retest reliability as well as statistically-significant known groups validity and/or responsiveness to change. Rather than striving for a one-size-fits-all approach to clinical validation, we have shown that certain digital measures hold utility for certain applications: measures that differentiate between PwPD and HCs (such as tap regularity) show potential as screening tools to aid clinical trial recruitment; measures that respond to symptomatic treatment (such as gait speed) may be useful in clinical care or in the development of novel symptomatic treatment medications; and measures that detect within-individual longitudinal disease progression (such as RMS acceleration during postural tremor) can be useful in the development of novel disease-modifying therapies. We have also shown that our digital measures are more reliable when summarized across 7-day bursts rather than individual assessments, though this might come at a cost of power for measuring progression.^15^ Together this emphasizes the many considerations that contribute to study design decisions.

We posit that all measures applied in research or clinical care should be reliable regardless of the CoU, and therefore the 22 out of 31 measures with test-retest reliability ICCs >0.5 are of primary interest (**Figure 4**). Some might argue that measures should also be required to exhibit criterion validity to be considered useful, but that assumes that the legacy measure used for assessing criterion validity does not have its own limitations (e.g., floor effects, lack of sensitivity, not being comprehensive, and lack of relevance to patients).^20–22^ Furthermore although the digital measures capture the same concept of interest as their aligned MDS-UPDRS item(s), the measures themselves are not directly comparable. For example, MDS-UPDRS item 2.10 (patient-reported *Tremor*) describes the impact of tremor on activities of daily living while item 3.15 (clinician-reported *Postural Tremor of the Hands*) describes tremor amplitude quantified by the human eye, neither of which would necessarily be expected to correlate perfectly with sensor-derived measures of acceleration and displacement. There is also the issue of multidimensional assessments such as MDS-UPDRS item 3.4 (*Finger Tapping*) which instructs the clinician to consider speed, amplitude, hesitations, halts, and decrementing amplitude when determining their score on a scale of 0-4. A high score may therefore be driven disproportionately by just one of these characteristics, thereby resulting in varying associations with individual digital measures of tapping. Finally, the accuracy and precision of sensors are likely to detect characteristics of motor function that are not readily observable, such as step time discrepancy or stride similarity, but which can be rigorously evaluated during analytical validation against an appropriate reference measure.^23^ Thus, we view criterion validity as helpful but not always necessary, and encourage investigators to explore other methods of demonstrating that a particular digital measure appropriately reflects the concept of interest.^24^

Clinical validity of a measure is also driven by how data is collected. Like many prior studies, ^4,5,25,26^ we were able to show that unsupervised both *in-home* administered assessments showed validity in addition to in-clinic supervised assessments (**Figure 2, supplemental Table 2**) this is in contrast to the Watch-PD study which were only able to demonstrate validity for the in-clinic assessments.^6,7^ In line with this study however, our analysis of the in-clinic measures tended to have stronger correlations for the criterion validity; in contrast, we had more statistically significant measures from the at-home measures **(supplemental Table 2)**. This can in part be explained by increased power and better estimates of average performance in the at-home measures where we had 278 data points that were obtained by averaging over 20 days of performance versus 102 for in-clinic which were derived from single administrations in the clinic.

A lack of power may also explain why we saw so few measures (6/31) being significant for known group validity analysis (as compared with 20 out of 31 for the response to medication, for example). This study is relatively small when it comes to the number of individuals (n=46) but large in the number of repeated measures (32,056) with hundreds of measures before and after medication for each PwPD. For known group validity it is the former that matters, something that is also made worse by the large age imbalance between the cohorts (61 years in PwPD versus 43 years in HC, on average). Furthermore, with individual variation generally being large for digital health data^27^ modeling within individual changes can be easier than finding across participant differences. A much larger and better matched dataset is required to build a diagnostic biomarker, as was demonstrated previously. ^28^

Both Watch-PD^7^ and At-Home-PD^17,29^ have shown that progression can be complicated by natural changes in performance that are unrelated to disease progression. Both of these studies, for example, reported measures of finger tapping improved with time in PwPD and, like here, Watch-PD reported improvements in HC participants as well. This may be due to learning or training effects, changes in participants’ compliance with specific assessment instructions over a long study, and general aging effects unrelated to disease progression. We chose to evaluate differential disease progression between PwPD versus HCs, rather than simple progression in the PwPD population, to mitigate the possibility of erroneously attributing changes with time to disease progression. To evaluate this choice we also assessed the changes in performance for the 31 measures in the HC cohort and found that 10/31 measures significantly changed. We realize that the relative progression is not the ideal design for a clinical study and suggest more work needs to be done to understand natural changes in performance to inform both study design and analysis. This could include better training and baseline data to avoid learning, modeling that accounts for a learning phase such as non-linear regression or collection of norming data to be used in analysis.

Our results should be interpreted alongside several limitations. This work is a re-analysis of existing data; hence many of the results presented in our figures are based on *p*-values obtained from tests that were not statistically powered *a priori*. Additionally, our analyses focused on PD motor symptoms captured during the assessments completed during the ObjectivePD substudy, and we were therefore not able to evaluate measures derived from other active assessments such as speech tasks or pronation/supination of the forearm, or from passively-captured data not tied to completion of timed assessments.^7,14,30,31^ Relatedly, although Supplementary Figure 1 depicts correlations between all measure combinations and therefore provides some insights into convergent and discriminant validity, the limited range of assessments which focused only on motor symptoms did not allow us to complete a thorough analysis of construct validity. Finally, although evaluating multiple aspects of clinical validity is a strength, it should be noted that our statistical analyses do not address theoretical aspects of measurement performance such as content validity, which is typically prioritized over other forms of validity described here. ^3,32^ and has been evaluated previously.^33^ Regardless of application, it is critical that all digital measures reflect a meaningful aspect of health and comprehensively capture the construct of interest.

By generating evidence to support various aspects of clinical validity in unsupervised settings, our study adds to the body of work suggesting that implementation of sDHTs can fulfill their promise of generating reliable and clinically-valid markers of disease state in decentralized trials and similar applications. The contrast between our context-driven approach and prior work underscores a key message: with appropriate methods of establishing clinical validity for specified CoUs, at-home digital measures can achieve clinical relevance on par with established endpoints, thereby bridging the gap between technological innovation and regulatory evidence.

## Data Availability

This study used publicly available, pseudonymized data. These data were contributed by users of the Parkinson mPower mobile application as part of the mPower study developed by Sage Bionetworks and described in Synapse [doi:10.7303/syn4993293].

https://www.synapse.org/mpower

## Acknowledgements

These data were contributed by users of the Parkinson mPower mobile application as part of the mPower study developed by Sage Bionetworks and described in Synapse [doi:10.7303/syn4993293].

## Supplemental Materials

**Supplementary Figure 1:**
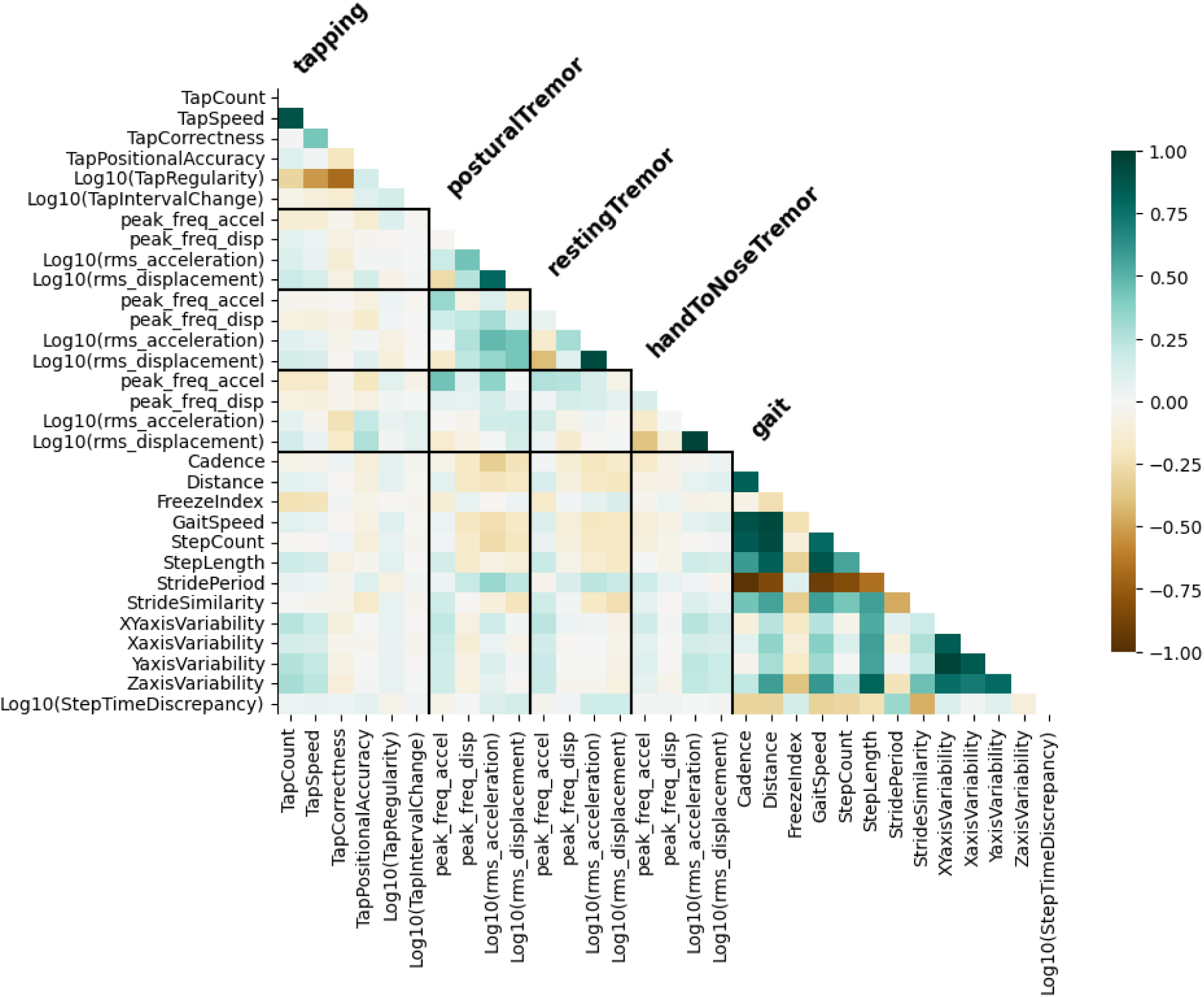
Correlation matrix including all digital measures organized by the assessment. Heatmap is colored by the Pearson Correlation between pairs of measures.

**Supplementary Table 1:**
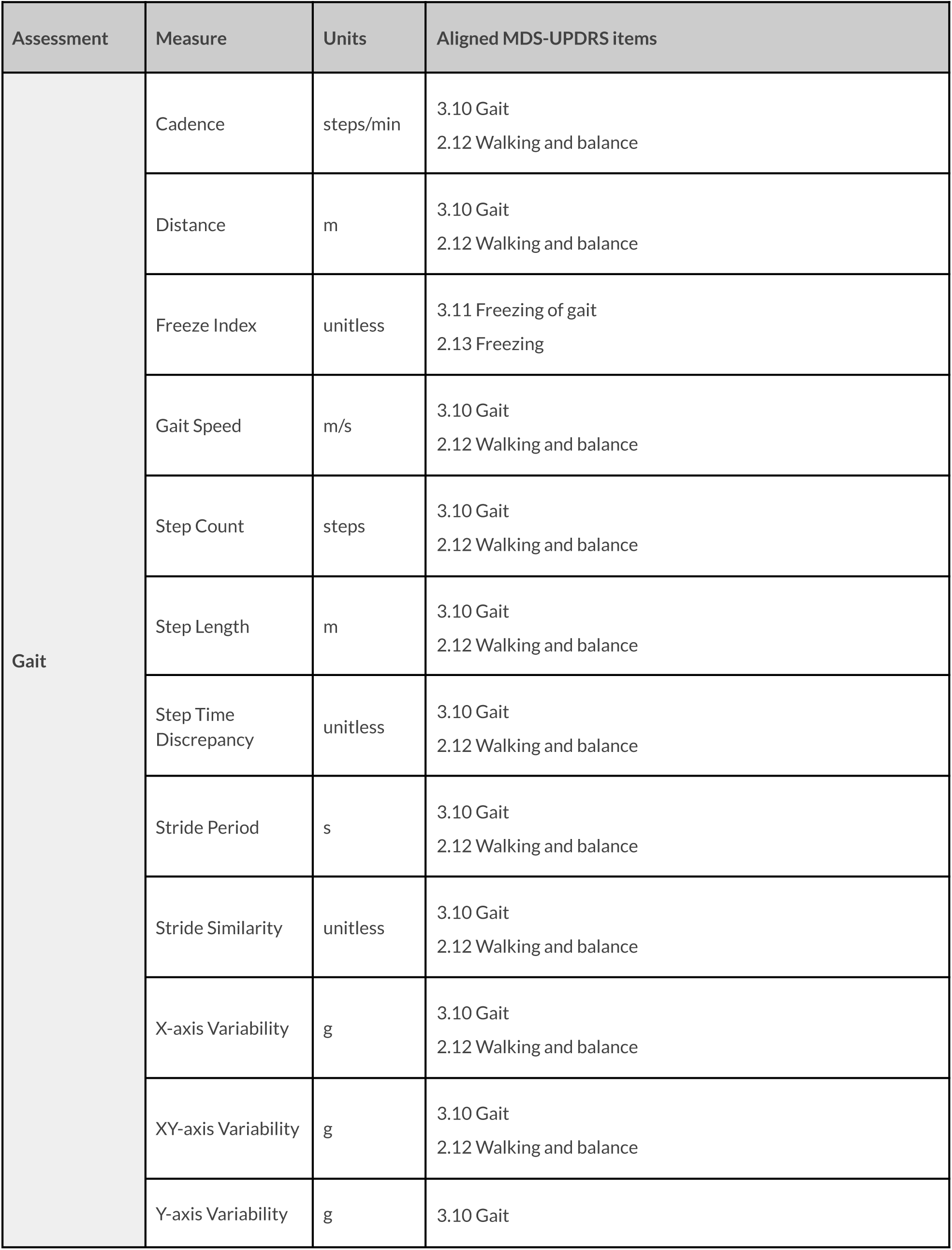
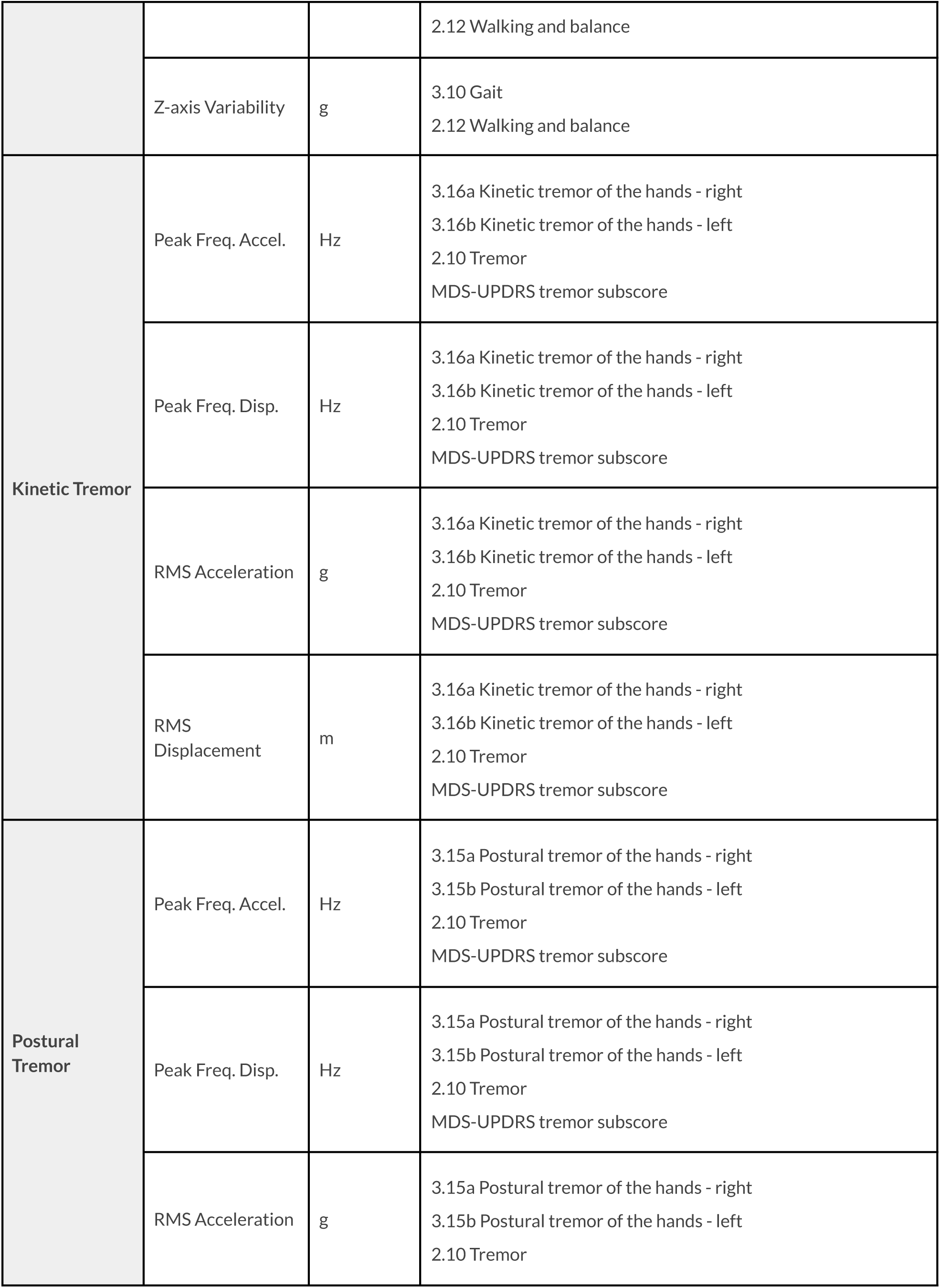
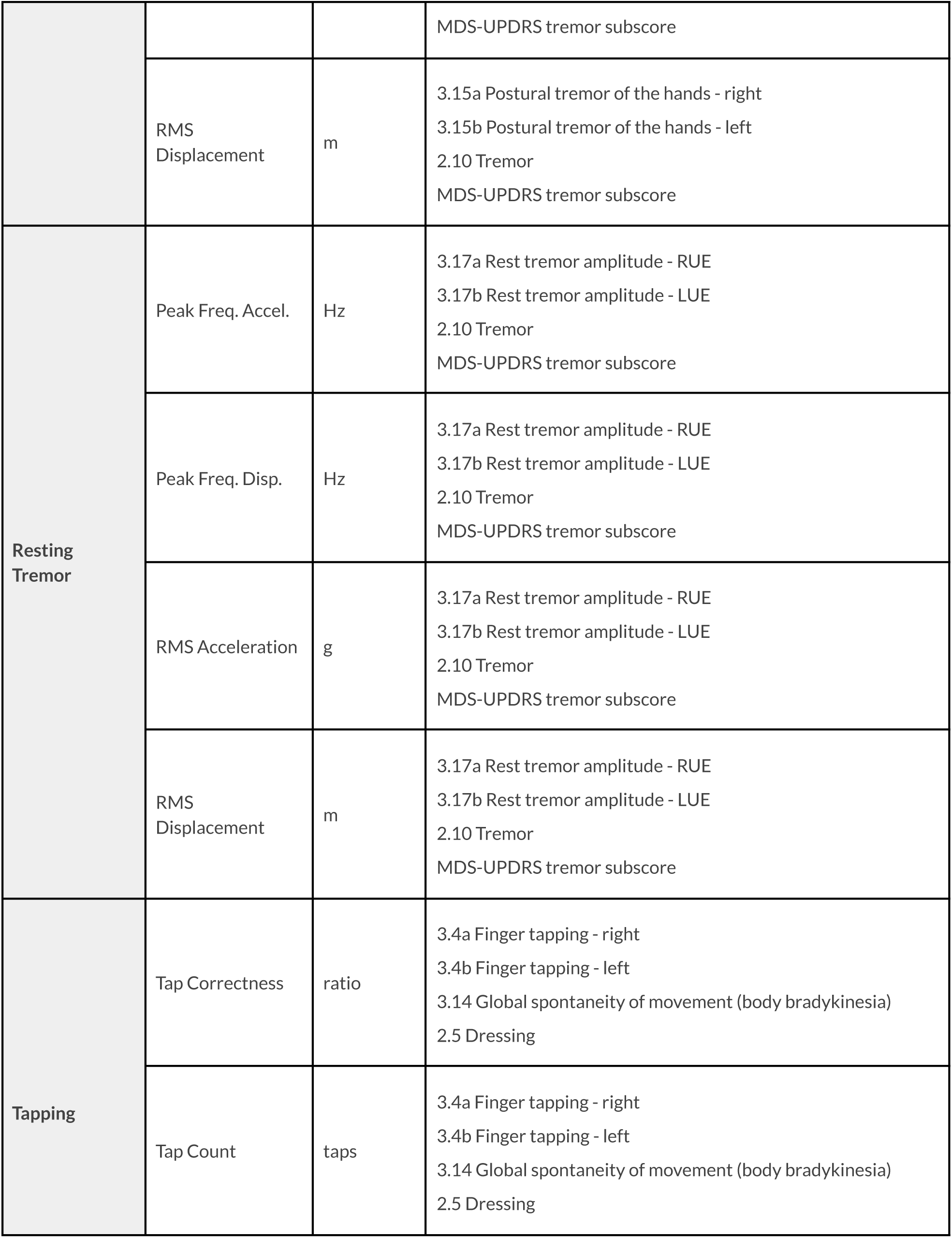
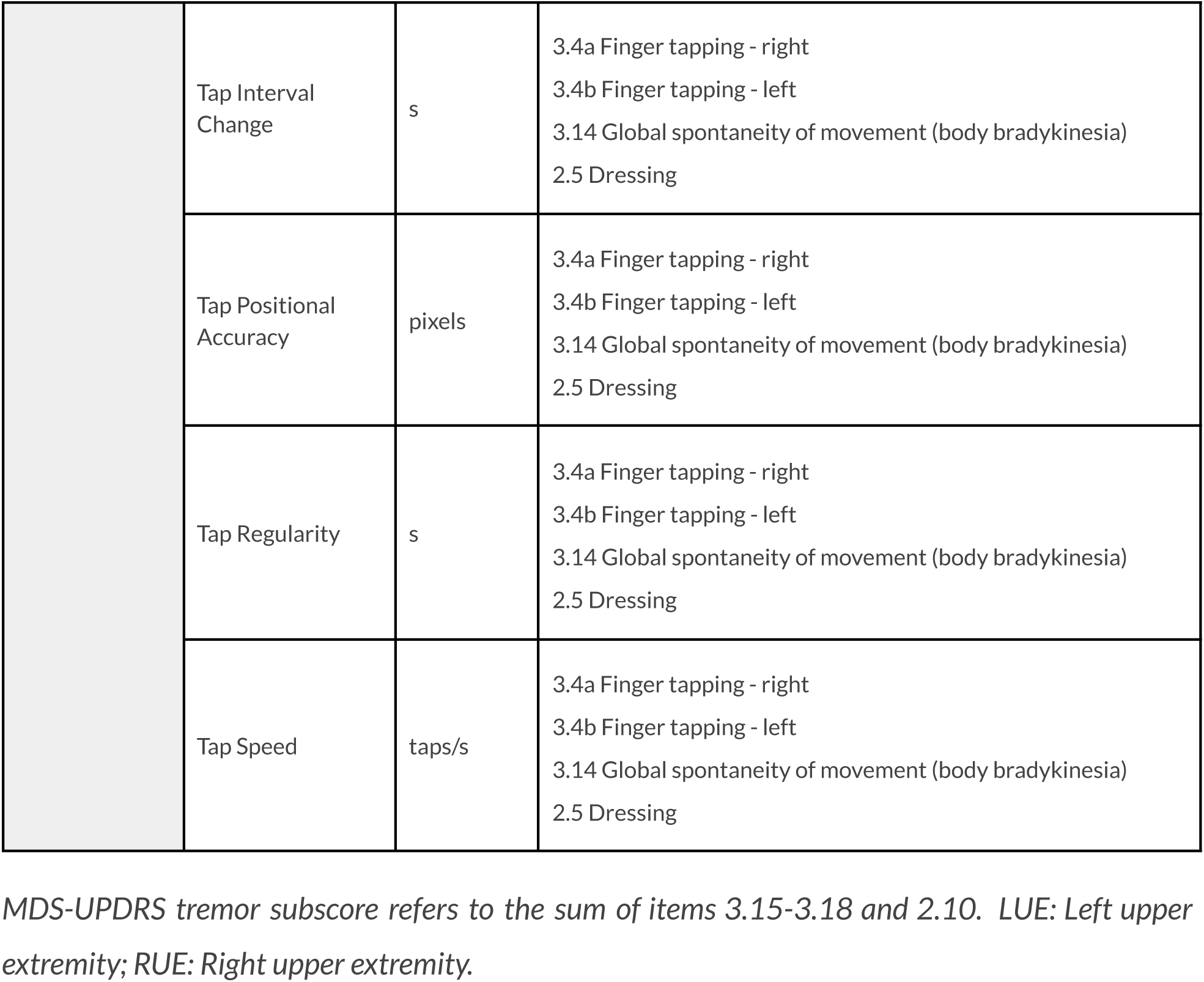
List of all digital measures and their aligned MDS-UPDRS items.

**Supplementary Table 2.**
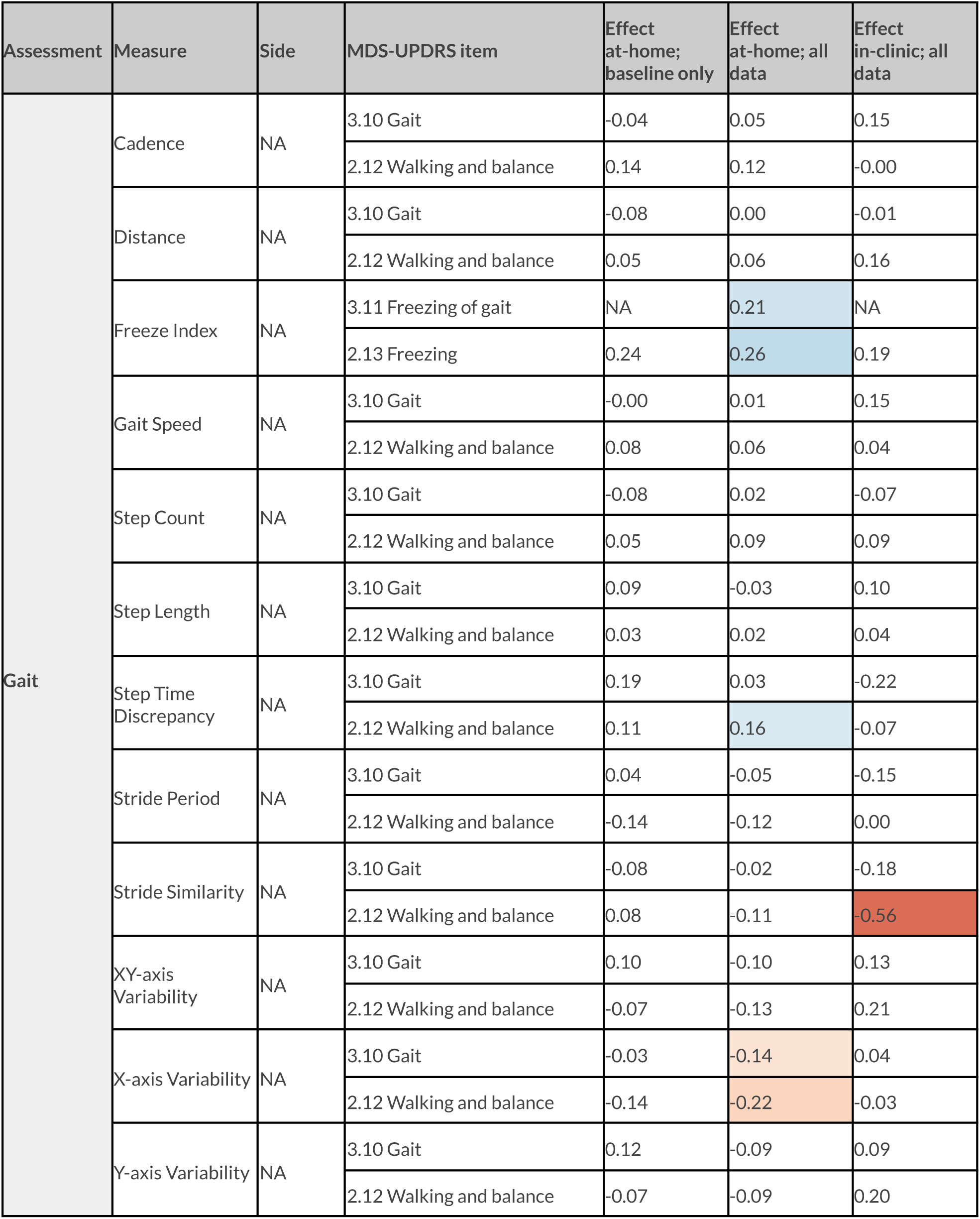
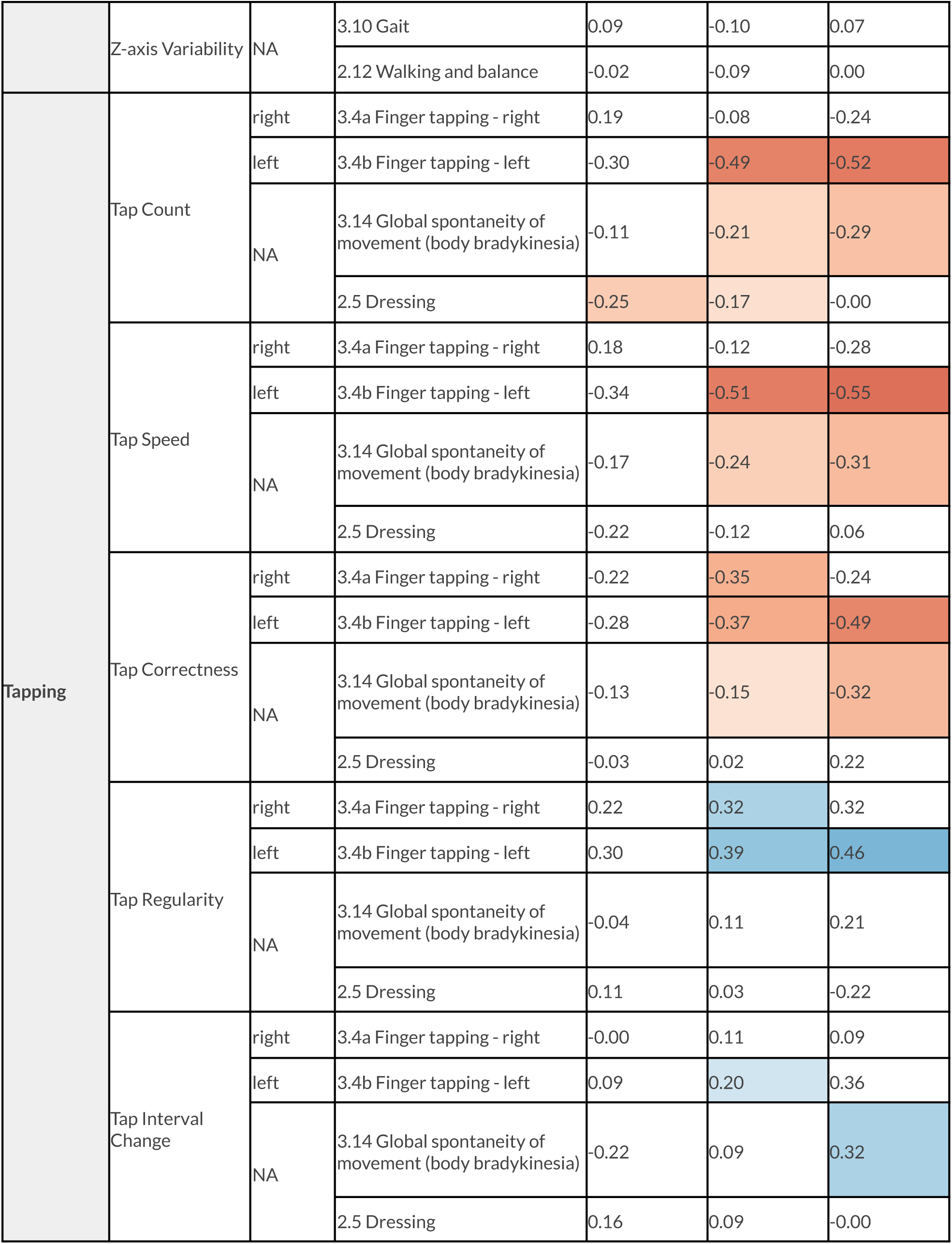
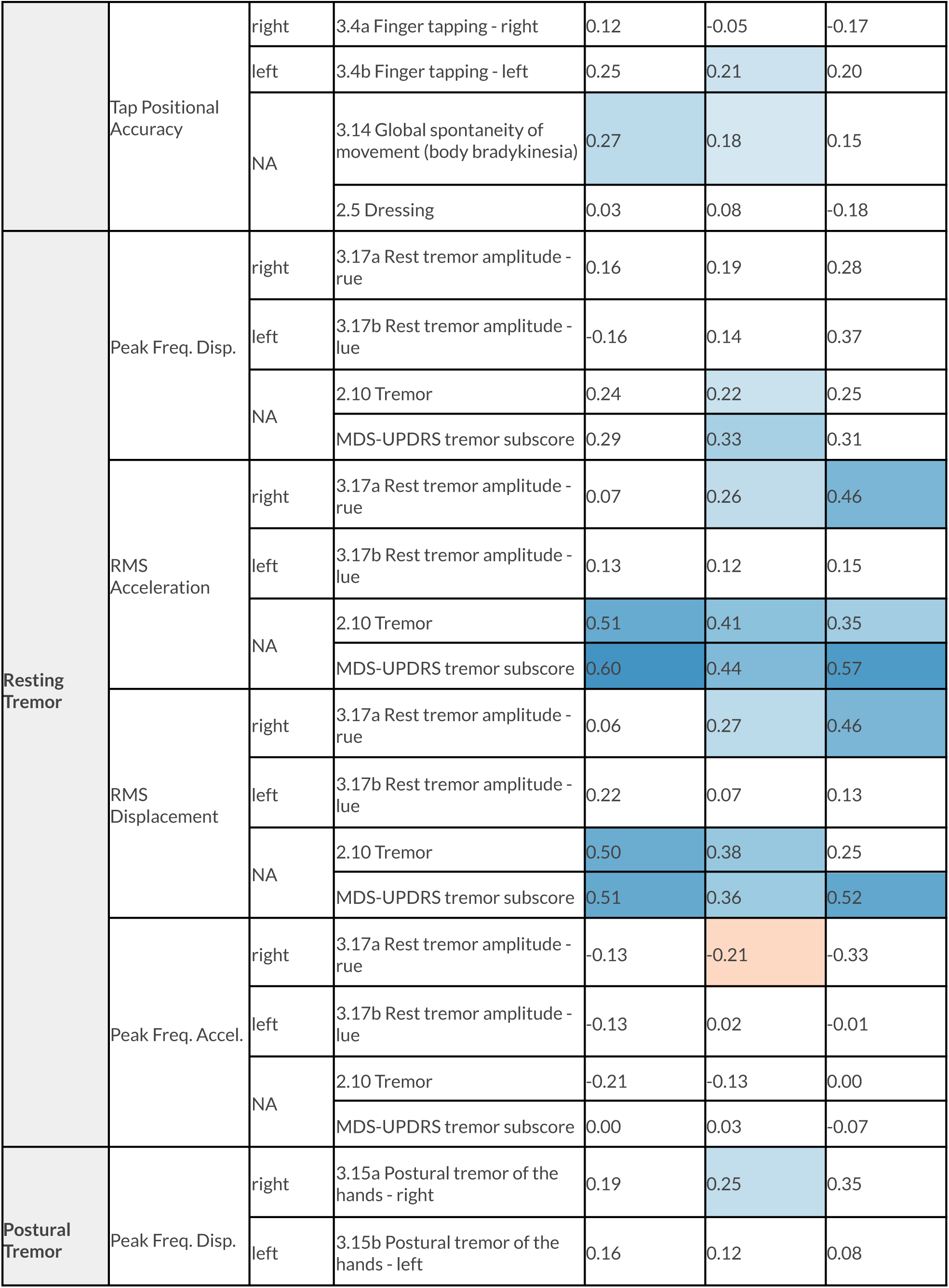
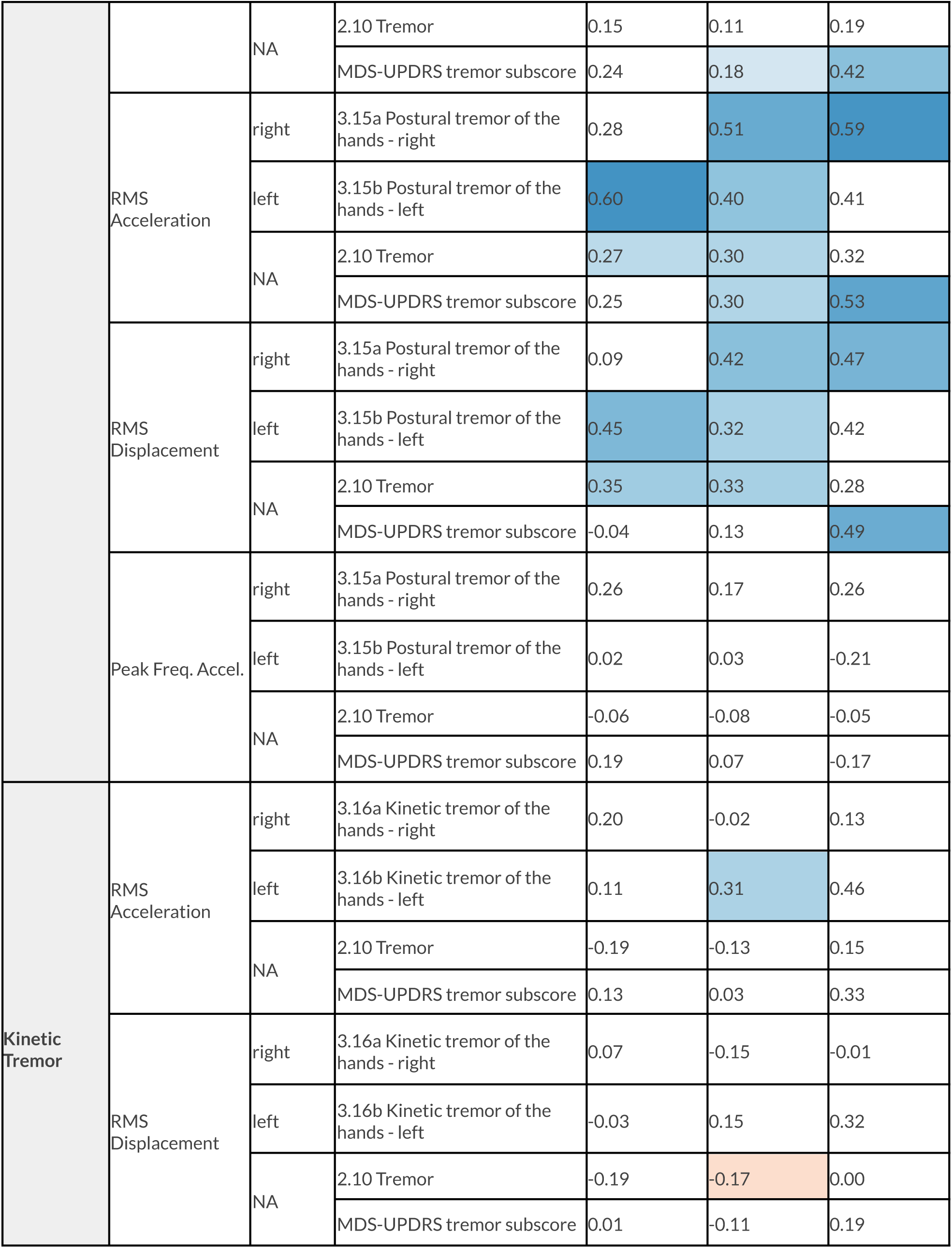
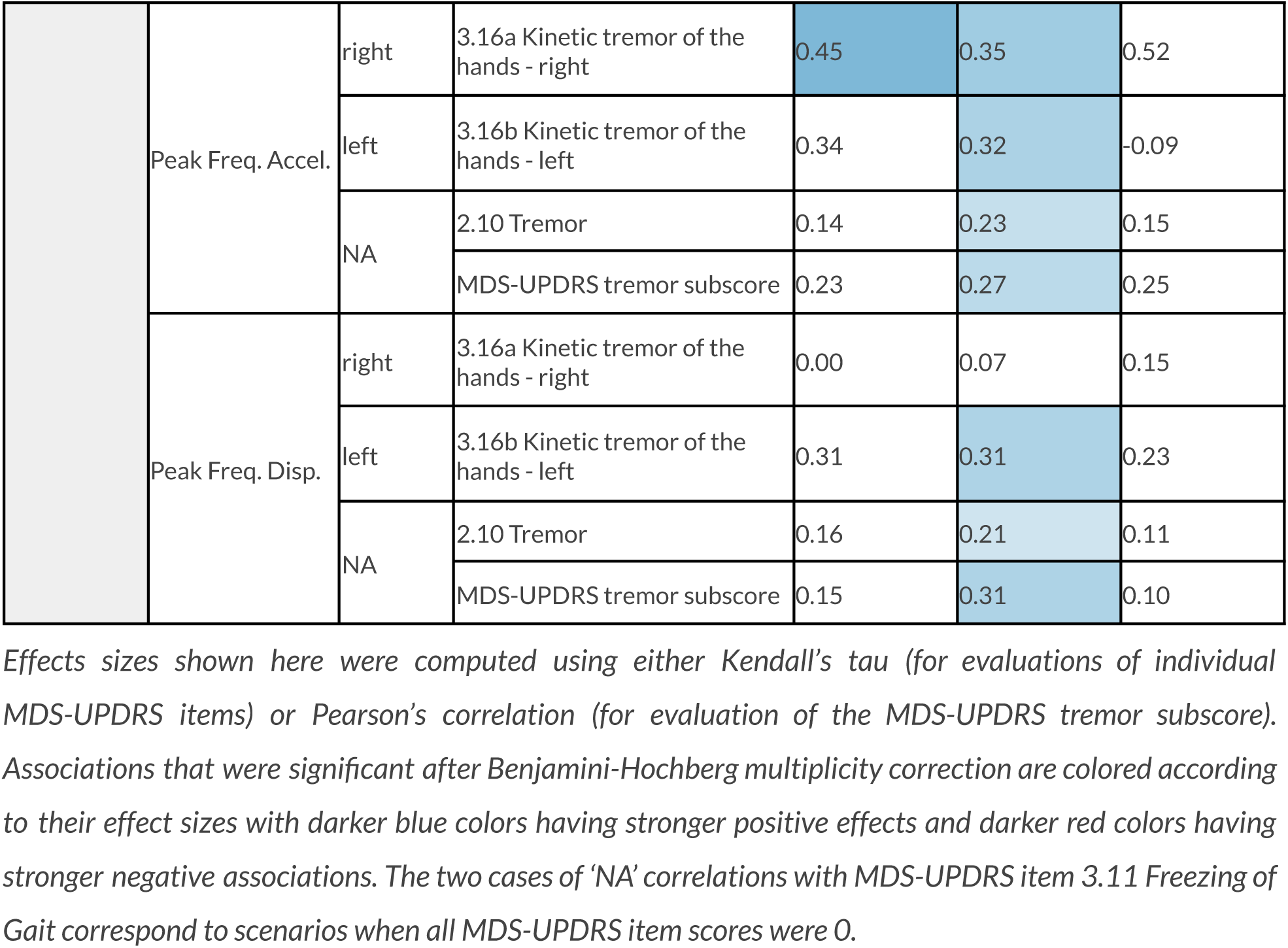
Associations between digital measures and MDS-UPDRS items by assessment timing and location.

